# Establishment and Quality Assessment of a Hospital-Associated Disability Database in Japan

**DOI:** 10.1101/2025.11.01.25339287

**Authors:** Shinsuke Hori, Hidetaka Wakabayashi, Shinta Nishioka, Takako Nagai, Eiji Kose, Nao Hashida, Kenta Ushida, Ryo Momosaki

## Abstract

**Objectives:** Hospital-associated disability (HAD) refers to a decline in activities of daily living (ADL) during hospitalization and is associated with prolonged hospital stay, higher mortality, and increased readmission rates among older adults. This study aimed to establish and assess the quality of a multicenter HAD registry database that includes detailed data on nutrition, medication, swallowing, and walking ability.

**Methods:** This prospective multicenter study was conducted at nine hospitals in Japan. Patients aged ≥70 years who were independent in ADL (Katz Index = 6) before admission to a general ward for rehabilitation were enrolled. Data were collected at admission and discharge using REDCap. Assessment items included demographics, Katz Index, Functional Comorbidity Index, Clinical Frailty Scale, Global Leadership Initiative on Malnutrition (GLIM) criteria, Food Intake LEVEL Scale (FILS), and Functional Ambulation Categories. HAD was defined as a Katz Index score of less than 6 on discharge.

**Results:** A total of 209 patients (median age = 79.0 [75–85] years; 60.8% male) were enrolled. The prevalence of HAD, hospital-associated dysphagia, and weight loss during hospitalization was 29.1%, 16.7%, and 75.4%, respectively. Median length of stay was 19 days (IQR: 13–34), and 82.3% were discharged home. While most variables had low levels of missing data, variables involving body weight demonstrated comparatively higher missingness.

**Conclusions:** A multicenter HAD registry database was successfully established, demonstrating the feasibility of prospective data collection across multiple institutions. This registry may facilitate future studies on risk factor identification, predictive model development, and early intervention strategies for HAD prevention.

## Introduction

Hospital-Associated Disability (HAD) refers to decline in Activities of Daily Living (ADL) that occurs during hospitalization.^1)^ Once patients develop HAD, they often fail to regain their pre-hospitalization level of function after discharge.^2)^ Decreases in ADL associated with hospitalization include mobility impairments and limitations in ADL, such as bathing, dressing, eating, and toileting.^3)^ Another functional impairment associated with hospitalization is hospital-associated dysphagia.^4,5)^ Hospital-associated dysphagia occurs as the patient’s general condition deteriorates during hospitalization, leading to a decline in swallowing ability and an increased risk of aspiration pneumonia.^6)^ HAD is particularly prevalent in older and frail patients and is associated with longer hospital stays, higher readmission prevalence and increased short- and long-term mortality.^7,8)^ In addition, the onset of HAD increases medical costs and the burden of care, posing a significant challenge to patients and to society as a whole.^9)^ Therefore, the need for HAD prevention and appropriate intervention is heightened.

Major risk factors for HAD include age, low activity during hospitalization, frailty, delirium, multiple medications, malnutrition, and prolonged hospitalization.^10,11)^ Reduced activity during hospitalization results in loss of muscle strength and physical fitness, making it challenging to perform ADL. Delirium is linked to reduced cognitive function and ADL, and it is known to negatively impact long-term prognosis. Moreover, polypharmacy increases the risk of adverse drug interactions and may contribute to the development of HAD. Furthermore, malnutrition after hospitalization can cause muscle weakness and inhibit recovery after discharge. Consequently, a variety of risks associated with HAD can be expected, and further research is necessary.

HAD, in a broad sense, refers to a decline in ADL related to hospitalization regardless of ADL status before hospitalization. On the other hand, narrowly defined HAD refers to the loss of independence in patients who were independent in ADL before hospitalization. Patients who were independent in ADL before hospitalization may require long-term care after losing their independence, which is a significant challenge in healthcare policy. The most critical point in conducting strict HAD research is accurately assessing the ADL status before hospitalization. However, previous databases often lack sufficient information on preadmission ADL, making it difficult to accurately identify HAD. Even prospectively collected databases have not accumulated detailed data on nutrition and medication use, ambulation, cognitive function, and oral intake ability.^12)^ The purpose of this study was to develop and evaluate the quality of a multicenter registry database on HAD, including detailed information on nutrition and medications, ambulation, and oral intake ability.

## Materials and Methods

Mie University Hospital served as the leading research institute, and partner facilities were recruited from the Japanese Society of Rehabilitation Nutrition. A total of nine facilities participated and prospectively constructed the database. The database employs a web-based registration system and was built using Research Electronic Data Capture (REDCap), software developed by Vanderbilt University.^13)^ The data registration period was from February 1, 2023, to March 31, 2025.

Patients included in the database were ≥70 years old, admitted to a general ward, and underwent rehabilitation. Eligible patients were required to have a diagnosis of a respiratory, cardiovascular, gastrointestinal, infectious, or malignant disease and a hospital stay of at least 48 hours. In addition, independence in ADL before admission was required, as assessed using the 6-item Katz Index.^14)^ The Katz Index is a tool used to evaluate the degree of independence in ADL among older adults or persons with disabilities, assessing six basic activities as either “independent (1 point)” or “dependent (0 points).” A total score of 6 points was considered indicative of independence in ADL. Preadmission Katz Index scores were determined by therapists based on information obtained from medical records, as well as from patients and/or their family members. Patients for whom preadmission ADL data were unavailable were not enrolled in the study.

To ensure data accuracy, manuals were provided to data entry personnel at each facility for entering patient data on admission and discharge. If the Katz Index on discharge was less than six points, HAD was considered to have developed. The study was approved by the Ethics Committee of Mie University Hospital (No. H2023-015), and informed consent was obtained from the participants in written form before registration.

### Assessment Items on Admission

Assessment items on admission include age, sex, type of the disease that led to admission, type of admission (emergency admission or scheduled admission), whether admitted by ambulance, stage of cancer modified Early Warning Score (MEWS),^15)^ use of ventilator support on admission, use of vasopressor drugs, consciousness status (Japan Coma Scale: JCS),^16)^ surgeries requiring general anesthesia, history of falls, and smoking status, as well as the Clinical Frailty Scale.^17)^ The MEWS is evaluated based on respiratory rate, heart rate, systolic blood pressure, level of consciousness, and temperature, with higher scores indicating greater severity. The JCS is a scoring system used to evaluate arousal status across four categories. Level 0 indicates full arousal, level 1 signifies arousal without stimulation, level 2 represents arousal with some stimulation, and level 3 denotes an inability to achieve arousal even with strong stimulation. The Clinical Frailty Scale is an index for evaluating frailty in older adults, consisting of nine levels ranging from 1 (very healthy) to 9 (terminal stage). It is based on a comprehensive evaluation of physical and cognitive functions. Additionally, Functional Comorbidity Index,^18)^ cognitive function score,^19)^ place of residence before admission (facility, hospital, home, other), medications used on admission (antipsychotics, anxiolytics, antidepressants, hypnotic–sedatives, antidementia medications, and number of medications), and laboratory values (albumin [Alb], C-reactive protein [CRP], hemoglobin [Hb]) on admission were recorded. The Functional Comorbidity Index is an index developed to assess comorbidities that affect physical function. It scores the presence of 13 chronic conditions, such as arthritis, osteoporosis, and depression, with higher total scores indicating a higher risk of physical decline. The cognitive function score assesses the level of independence among older adults with dementia. It features a ranking system from 0 to 5, where 0 indicates complete independence and 5 signifies significant mental symptoms and problematic behaviors that require specialized medical care. This index helps to reflect the need for assistance in daily living activities and the stability of the individual’s behavior. Additionally, the following information was recorded on admission: the Katz Index, the Food Intake LEVEL Scale (FILS),^20)^ Functional Ambulation Categories (FAC),^21)^ and Global Leadership Initiative on Malnutrition (GLIM) criteria.^22)^ Other recorded factors included height, weight, and weight from 3 to 6 months prior as well as the need for infection control in rehabilitation implementation, bed rest orders, urinary catheter placement, and the use of physical restraints. The FILS is a rating scale for eating and swallowing abilities, consisting of 10 steps from 1 (unable to eat food orally) to 10 (able to eat normal food), which indicates the current status of oral intake. The FAC is a walking ability rating scale that categorizes walking capability on a 6-point scale from 0 (unable to walk) to 5 (independent walking), based on the level of assistance required for walking. The GLIM criteria are internationally standardized criteria for diagnosing malnutrition. The diagnosis using the GLIM criteria is a two-step process. First, patients are identified using a validated screening tool. To be diagnosed as malnourished, they must meet at least one criterion from the phenotypic category, which includes weight loss, low body mass index (BMI), and muscle mass loss, and the etiologic category, which includes factors such as inflammation, disease burden, and insufficient intake. Cases diagnosed as malnourished were considered severely malnourished if any one of the three phenotypic criteria exceeds the threshold for severe malnutrition; otherwise, they are categorized as moderately malnourished. Laboratory values were obtained within 3 days of admission using the values closest to the admission date.

### Discharge Assessment Items

Items evaluated on discharge include the length of stay and the discharge destination, which can be a general ward, convalescent ward, long-term care ward, home, facility, or death. Additionally, the assessment covers any complications experienced during hospitalization, such as pneumonia, falls, depression, or delirium. It also includes whether physical therapy, occupational therapy, or speech therapy was provided, along with the total number of therapy units administered (1 unit = 20 minutes). Additionally, the FILS score on discharge and the rehabilitation start location (bedside or training room), as well as the Katz Index and FAC on discharge, were also recorded. The presence of dental treatments, whether the patient is edentulous, denture use, weight on discharge, length of nothing by mouth period after admission, infectious diseases during hospitalization, presence of parenteral nutrition, presence of enteral nutrition, medications and number of medication types administered on discharge were also recorded. HAD was defined as a Katz Index score < 6 on discharge. Hospital-associated dysphagia was defined as a decrease in FILS after admission (Food Intake LEVEL Scale on discharge < Food Intake LEVEL Scale on admission).

Additionally, facility Information: the number of general hospital beds, hospital type, total number of therapists, number of patients receiving rehabilitation, whether weekend rehabilitation was offered, and the availability of dentists and dental hygienists, number of full-time dietitians and ward dietitians at each facility was also recorded. Facility information was collected from each participating institution at the time of enrollment.

### Data Quality Assurance

The data dictionary is provided as a supplementary table. The data capture system was developed by the Osaka Metropolitan University REDCap Group. For ordinal variables, values outside the defined range cannot be selected. For continuous variables, outliers cannot be entered. A brief data entry manual was distributed to all data registrants. Each facility’s database manager was asked to monitor eligibility and case registration status within REDCap on an ongoing basis. Data entry was scheduled at two time points: admission and discharge. Clinical variables were entered based on physicians’ assessments; rehabilitation-related variables based on evaluations by rehabilitation professionals; and nutrition-related variables based on the judgment of ward staff or registered dietitians. The diagnosis of the condition leading to admission and comorbidities was based on physicians’ clinical judgment. For items that might be prone to ambiguity, explanatory notes were included in the data entry sheet. In particular, visual reference materials outlining the assessment criteria were provided for the Clinical Frailty Scale, the cognitive function score, the Katz Index, FILS, FAC, and the GLIM criteria. For patients who died during hospitalization, we requested that their clinical status immediately prior to death be recorded. In addition, the corresponding author reviewed the REDCap database monthly to identify outliers or missing values in required fields and requested clarification or data completion from participating sites when necessary.

### Statistical Analysis

Descriptive statistics were used to analyze the data. Frequencies and percentages were reported for categorical variables. As this study was descriptive in nature, no statistical hypothesis testing was conducted. IBM SPSS ver. 26 (IBM Corporation; Armonk, New York, US) was used for statistical analysis. Nominal data were expressed as numbers and percentages (%), whereas continuous data were reported as mean ± standard deviation (SD) for parametric values and as median (interquartile range [IQR]: 25th–75th percentile) for nonparametric values assessed via histogram. The number of missing values was also recorded.

We conducted an additional facility-level comparative analysis limited to the facilities that enrolled ≥10 patients. Facility-level analyses were performed for the main outcome, HAD, and for missingness in GLIM-defined malnutrition.

This article was previously posted as a preprint on medRxiv (DOI: https://doi.org/10.1101/2025.11.01.25339287) on Nov 01, 2025. This work is licensed under a CC BY 4.0 License.

## Results

Among the nine hospitals involved in the HAD database, the total number of registered patients was 209. Table 1 displays the data recorded on admission. Variables with missing values exceeding 5% included cancer stage (13.3%), BMI on admission (5.0%), malnutrition on admission (20.6%), weight 3–6 months before admission (41.6%), and average energy intake within 48 hours after admission (5.0%). The median age of patients in the study was 79.0 [75–85] years, with 60.8% of participants being men. Cancers accounted for 39.7% of hospitalizations, followed by cardiovascular diseases at 29.7% and respiratory diseases at 15.3%. In terms of hospitalization type, 52.2% were scheduled admissions, 47.8% were emergency admissions, and 28% involved ambulance transports. Among patients with cancer, 28.9% were Stage 3 and 20.5% were Stage 4.

**Table. 1.** Admission data (n=209)

| | Number of<br>missing (%) | n (%), mean ± SD,<br>median [IQR] |
| --- | --- | --- |
| Age, years | 0 | 79.0 [75–85] |
| Sex, Male | 0 | 127 (60.8) |
| Primary disease leading to hospitalization | 0 |  |
| Malignant tumor |  | 83 (39.7) |
| Circulatory disease |  | 62 (29.7) |
| Gastrointestinal disease |  | 13 (6.2) |
| Infectious disease |  | 19 (9.1) |
| Respiratory disease |  | 32 (15.3) |
| Hospitalization type | 0 |  |
| Scheduled hospitalization |  | 109 (52.2) |
| Emergency hospitalization |  | 100 (47.8) |
| Place of residence before admission | 0 |  |
| General ward |  | 3 (1.4) |
| Home |  | 199 (95.2) |
| Facility |  | 7 (3.3) |
| Stage of Cancer (among 83 patients with cancer) | 11 (13.3) |  |
| 0 |  | 3 (3.6) |
| 1 |  | 14 (16.9) |
| 2 |  | 14 (16.9) |
| 3 |  | 24 (28.9) |
| 4 |  | 17 (20.5) |
| Admission via ambulance | 2 (1) | 58 (28) |
| Modified early warning score on admission | 0 | 0.0 [0.0–2.0] |
| Japan Coma Scale on admission | 0 |  |
| 0 |  | 197 (94.3) |
| 1 |  | 8 (3.8) |
| 2 |  | 3 (1.4) |
| 3 |  | 1 (0.5) |
| Vasopressors use on admission | 2 (1) | 5 (2.4) |
| Ventilator use on admission | 0 | 20 (9.6) |
| Surgery with general anesthesia during hospitalization | 0 | 64 (30.6) |
| History of falls in the year before admission | 0 | 19 (9.1) |
| Smoking habits in the year before admission | 1 (0.5) | 33 (15.8) |
| Cognitive function score before hospitalization | 0 |  |
| 0 |  | 148 (70.8) |
| 1 |  | 42 (20.1) |
| 2 |  | 15 (7.2) |
| 3 |  | 3 (1.4) |
| 4 |  | 1 (0.5) |
| Preadmission Clinical Frailty Scale | 1 (0.5) |  |
| 1 |  | 27 (13) |
| 2 |  | 34 (16.3) |
| 3 |  | 61 (29.3) |
| 4 |  | 40 (19.2) |
| 5 |  | 46 (22.1) |
| Functional comorbidity index on admission | 0 |  |
| 0 |  | 51 (24.4) |
| 1 |  | 75 (35.9) |
| 2 |  | 50 (23.9) |
| 3 |  | 23 (11.0) |
| 4 |  | 6 (2.9) |
| 5 |  | 4 (1.9) |
| Number of drug types on admission | 5 (2.4) | 5.5 [3.0–8.0] |
| Antipsychotics on admission | 1 (0.5) | 5 (2.4) |
| Antianxiety medications on admission | 1 (0.5) | 7 (3.3) |
| Antidepressants on admission | 1 (0.5) | 5 (2.4) |
| Hypnotic–sedatives on admission | 1 (0.5) | 27 (12.9) |
| Antidementia medications on admission | 1 (0.5) | 8 (3.8) |
| Alb on admission, g/dL | 5 (2.4) | 3.5 [3.1–3.9] |
| CRP on admission, mg/dL | 8 (3.8) | 0.9 [0.1–6.1] |
| Hb on admission, g/dL | 3 (1.4) | 11.6 [10.2–12.9] |
| Katz Index on admission | 0 | 6.0 [5.0–6.0] |
| Food Intake LEVEL Scale on admission | 2 (1) |  |
| No oral intake (1–3) |  | 17 (8.2) |
| Oral intake and alternative nutrition (4–6) |  | 8 (3.9) |
| Oral intake only (7–9) |  | 47 (22.7) |
| Normal (10) |  | 135 (65.2) |
| Functional Ambulation Categories on admission | 1 (0.5) |  |
| 0 |  | 26 (12.5) |
| 1 |  | 7 (3.4) |
| 2 |  | 6 (2.9) |
| 3 |  | 22 (10.6) |
| 4 |  | 49 (23.6) |
| 5 |  | 98 (47.1) |
| Diagnosis of malnutrition on admission (GLIM criteria) | 43 (20.6) |  |
| No malnutrition |  | 99 (59.6) |
| Moderate malnutrition |  | 37 (22.3) |
| Severe malnutrition |  | 30 (18.1) |
| Weight on admission, kg | 7 (3.3) | 55.2 [47.9–63.7] |
| Body mass index on admission | 10 (5) | 21.9 [19.7–24.5] |
| Weight 3–6 months before admission, kg | 87 (41.6) | 57.0 [48.0–65.0] |
| Average energy intake within 48 hours of admission, kcal | 10 (5) | 1400 [800–1600] |
| Bed rest order on admission | 2 (1) | 35 (16.9) |
| Need for infection control in rehabilitation implementation | 1 (0.5) | 4 (1.9) |
| Urinary catheter placement on admission | 3 (1.4) | 36 (17.5) |
| Physical restraint on admission | 2 (1) | 7 (3.4) |

On admission, the clinical status included a median MEWS of 0.0 [0.0–2.0], a JCS score of 0 for the majority (94.3%), use of a ventilator in 9.6% of cases, and administration of vasopressors in 2.4% of patients. Moreover, 30.6% had received surgery under general anesthesia, 9.1% had a history of falls, and 15.8% of the patients had a history of smoking. Regarding preadmission frailty indicators, 70.6% of patients had a Clinical Frailty Scale score of 3 or higher. The Functional Comorbidity Index had the highest score of 1 in 35.9% of patients. On admission, 12.9% of patients used hypnotic–sedatives, 3.8% used antidementia drugs, and <3% used antipsychotics, anxiolytics, or antidepressants. The median Katz Index on admission was 6 [5–6]. Among the patients, 65.2% had normal oral intake, whereas 12.1% were considered difficult to feed orally (FILS 1–6). The largest proportion of patients (47.1%) had a FAC score of 5, indicating independent walking, whereas 12.5% of patients were classified as FAC 0, meaning they were unable to walk. On admission, nutrition-related indices indicated a median BMI of 21.9 [19.7–24.5] and a median weight of 55.2 [47.9–63.7] kg over the previous 3–6 months. The average energy intake during the 48 hours following admission was 1400 [800–1600] kcal. According to the GLIM criteria for diagnosing malnutrition, 59.6% of patients were not malnourished, 22.3% had moderate malnutrition, and 18.1% had severe malnutrition. Additionally, 16.9% were on bed rest, 17.5% had urinary catheters, 3.4% underwent physical restraint, and 1.9% required infection control during rehabilitation.

Table 2 presents the data recorded on discharge. The three variables with missing values that exceeded 5% were BMI on discharge (15.3%), the length of nothing by mouth period (9.1%), and weight loss after admission (14.3%). The prevalence of HAD was 29.1%, whereas the prevalence of hospital-associated dysphagia was 16.7%. During hospitalization, 75.4% of patients experienced weight loss, and 15.9% decreased FAC score. The median length of hospital stay was 19.0 [13–34] days. The most common discharge destination was home (82.3%), followed by general wards (6.2%) and convalescent wards (5.3%). During hospitalization, adverse events included pneumonia (8.6%), falls (6.3%), depression (1.0%), and delirium (9.6%). Physical therapist intervention was present in 97.6% of cases, whereas occupational therapists were involved in 25.8%, and speech therapists in 14.4%. The median total number of rehabilitation units was 16.5 [8.0–31.0]. Swallowing ability on discharge based on FILS was as follows: 6.7% without oral intake, 4.3% with oral intake plus alternative nutrition, 28.2% with oral intake only, and 59.8% normal. Rehabilitation was initiated at the bedside in 72.7% of cases and in the training room in 27.3%. The median Katz Index on discharge was 5.0 [5.0–6.0]. For walking ability (FAC), 47.6% scored 5 and 26.9% scored 4. Cognitive function score was 63.8% for rank 0, 22.7% for rank 1, and 8.7% for rank 2, indicating that most patients maintained relatively high levels of independence. During hospitalization, 17.9% of patients received dental interventions. Of the patients, 8.8% were edentulous, 46.1% used dentures. The median body weight on discharge was 53.1 [45.4–61.3], with a missing data rate of 13.9%. The median length of nothing by mouth period after admission was 0 days [0–1]. Dietitians were involved in 50.7% of cases. Total parenteral nutrition was administered to 16.3% of patients, whereas enteral nutrition was given to 12.0%. The median number of drug types on discharge was 6 [3–8]. Usage of medications were as follows: antipsychotics 2.4%, anxiolytics 3.8%, antidepressants 2.4%, hypnotic–sedatives 20.2%, and antidementia drugs 2.9%.

**Table. 2.**
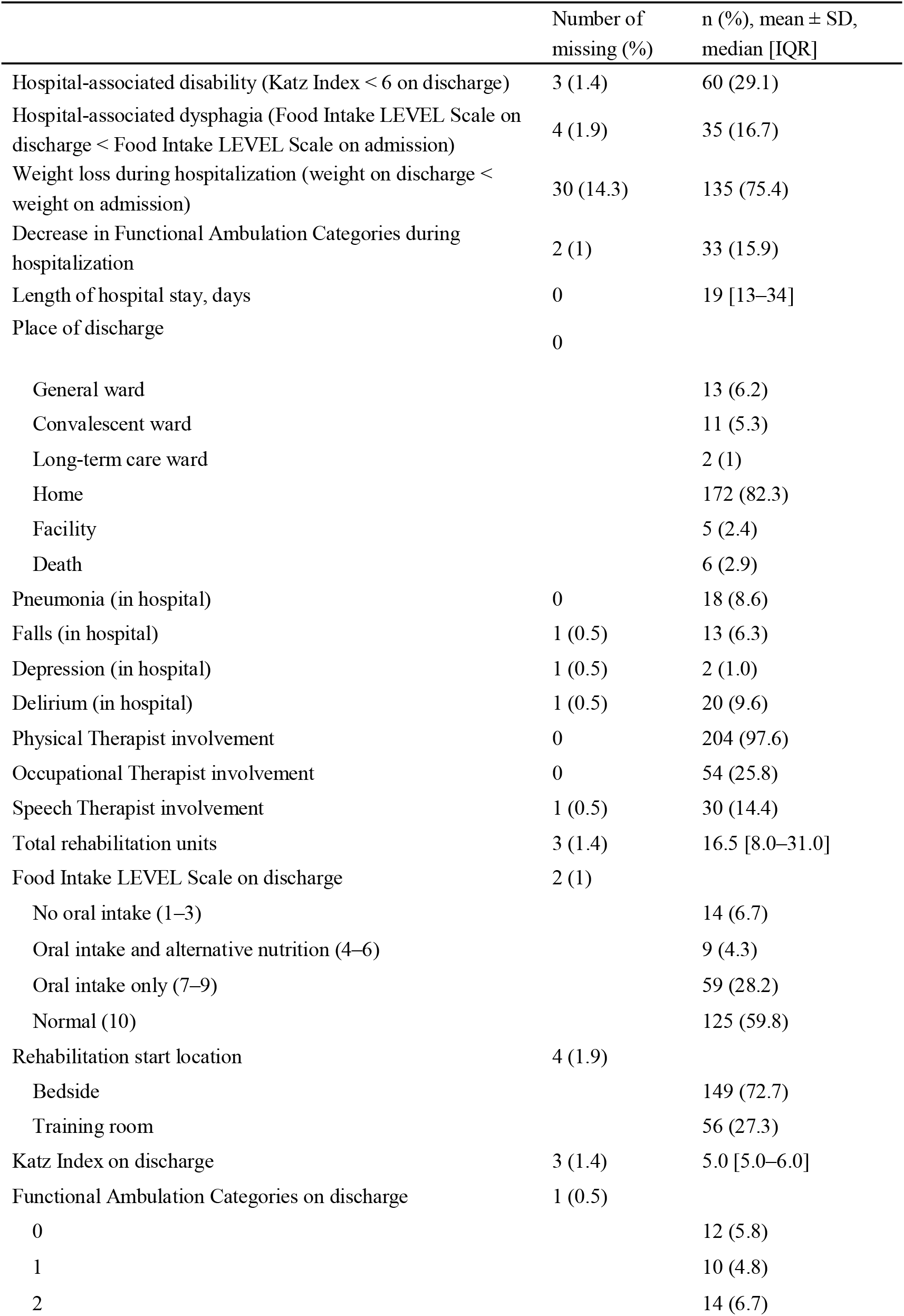
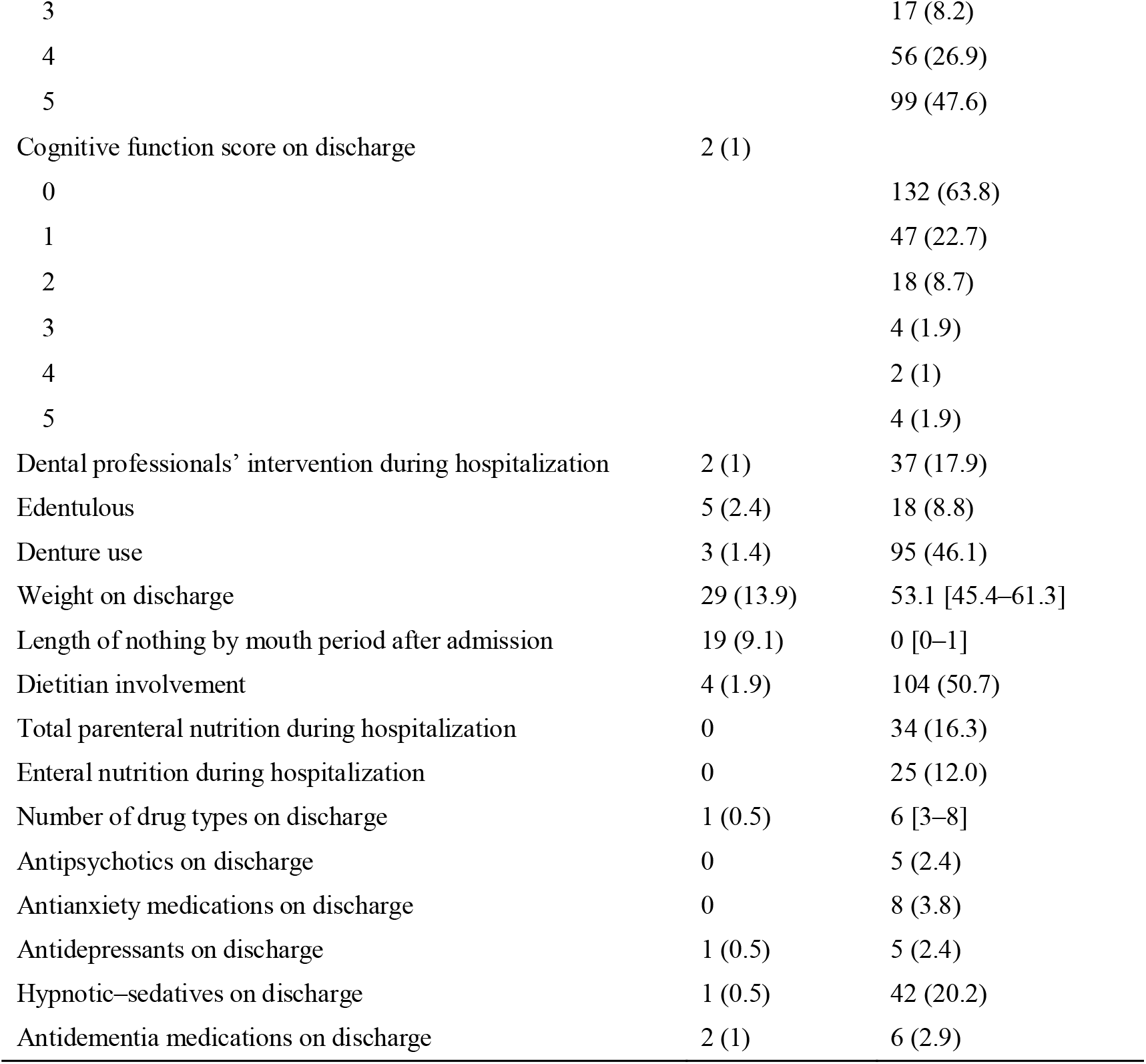
Discharge Data (n=209)

Table 3 contains information about the participating facilities. University hospitals made up 44.4% of the total. The average number of general beds was 472.3 (±218.5), and the median number of therapists was 41 [24–61]. All facilities had systems for holiday rehabilitation. The median number of ward dietitians was 1.0 [0.5–7.5]. The median number of registered patients per facility was 17 [11–26], with a substantial proportion of cases enrolled from general hospitals, including university hospitals. The minimum and maximum numbers of registered patients per facility were 1 and 88, respectively.

**Table. 3.** Facility Information (n=9)

| | Number of missing (%) | n (%), mean ± SD, median [IQR] |
| --- | --- | --- |
| University hospital | 0 | 3 (33.3) |
| Number of general hospital beds | 0 | 472.3 ± 218.5 |
| Total number of therapists | 0 | 41 [24–61] |
| Weekend rehabilitation | 0 | 9 (100) |
| Dentists | 0 | 5 (55.5) |
| Dental hygienists | 0 | 5 (55.5) |
| Number of ward dietitians | 0 | 1 [0.5–7.5] |

Table 4 shows an additional facility-level comparative analysis limited to the seven facilities that enrolled ≥10 patients. Facility-level analyses were performed for the main outcome, HAD, and for GLIM-defined malnutrition, which had a high proportion of missing data. The incidence of HAD varied across facilities. However, given that some facilities had relatively small sample sizes, direct comparison of incidence rates between facilities is limited by statistical instability. Missingness in GLIM-defined malnutrition was concentrated in specific facilities.

**Table 4.** Facility-level comparative analysis.

| Hospital ID | 1 | 2 | 3 | 4 | 5 | 6 | 7 |
| --- | --- | --- | --- | --- | --- | --- | --- |
| Number of eligible patients | 11 | 11 | 17 | 88 | 30 | 20 | 26 |
| Hospital-associated disability, n (%) | 4 (36.4) | 0 (0) | 12 (70.6) | 13 (14.8) | 8 (26.7) | 2 (10.0) | 18 (69.2) |
| Missingness in GLIM-defined malnutrition, n (%) | 0 (0) | 0 (0) | 0 (0) | 29 (33) | 0 (0) | 2 (10.0) | 12 (46.2) |

## Discussion

We established a multicenter prospective database on HAD and collected detailed clinical and functional data on 209 hospitalized older adults. In this database, HAD occurred in 29.1% of patients. Hospital-associated dysphagia occurred in 16.7% of patients, and 75.4% of hospitalized patients experienced weight loss during hospitalization. Additionally, the FAC score decreased in 15.9% of patients during hospitalization. While most variables had low levels of missing data, variables involving body weight—such as GLIM-defined malnutrition, body weight before admission, weight on discharge, and weight loss during hospitalization —demonstrated comparatively higher missingness.

The incidence of HAD in this study was 29.1%, which was consistent with that reported in previous studies. The reported incidence of HAD varied among studies, likely due to differences in patients’ baseline functional levels, timing of assessment, and HAD definitions. Zaslavsky et al.^23)^ and Brown et al.^24)^ reported higher incidence in older adults hospitalized in acute care settings. They assessed ADL at multiple time points, from before admission to 1 month after discharge. Using the Modified Barthel Index, Zaslavsky et al. defined HAD as failure to regain pre-hospital functional status after discharge, reporting an incidence of approximately 44%. In a randomized controlled trial of cognitively intact older adults, Brown et al. reported a 43% incidence of HAD in the usual care group. However, unlike our study, these studies did not exclusively include patients who were independent in ADL before hospitalization, which may have influenced the observed incidence of HAD. Differences in assessment timing, whether pre- and post-hospitalization data were available; baseline ADL; age and cognitive function; length of stay; and whether rehabilitation was provided could all contribute to variations in the reported incidence of HAD.

In this study, hospital-associated dysphagia was observed in 16.7% of the patients. In the previous study by Maeda et al.,^25)^ hospital-associated dysphagia was reported in 4.3% of patients, which is lower than the incidence observed in the present study. Nagano et al.^26)^ reported that 5.6% of patients who already had dysphagia on admission experienced a further decline in their swallowing ability during hospitalization. In that study, the group with decreased swallowing ability showed significantly higher rates of malnutrition, insufficient energy intake, and longer nothing per mouth durations. The difference in definition of hospital-associated dysphagia may account for the discrepancy in incidence between our study and previous research. In our study, we defined hospital-associated dysphagia as a decrease in the FILS, whereas the previous study defined it based on a change in diet form. Our study also differed from those by Maeda et al. and Nagano et al. in patient characteristics, including age and reasons for hospitalization, which may have influenced the outcomes. In the report by Nagano et al., the Functional Oral Intake Scale (FOIS) was used to assess swallowing function, and decreased swallowing ability was defined as FOIS ≤5. Although the FOIS and FILS are assessment tools that evaluate actual feeding conditions, the FILS provides a more detailed assessment. This difference may also explain the variation in the incidence of hospital-associated dysphagia between our study and previous research. Swallowing ability is closely associated with overall muscle strength, as represented by the concept manuscript of sarcopenic dysphagia.^27)^ Reduced physical activity and malnutrition during hospitalization can lead to deterioration of general health and impaired swallowing function. In our study, a high incidence of weight loss during hospitalization (75.4%) was also observed, suggesting that the presence or progression of sarcopenia may have contributed to the decline in swallowing ability. Unlike previous studies, our study included patients who were undergoing rehabilitation. Patients requiring rehabilitation may be more susceptible to sarcopenic effects, making them more prone to developing hospital-associated dysphagia.

In this study, 75.4% of the patients experienced weight loss during hospitalization. In previous studies, 41.2%–45.5% of patients were reported to have lost weight during hospitalization,^28,29)^ which was lower than in the present study. Zannidi et al.^30)^ reported that 19% of patients lost 10% or more of their weight during their hospital stay, and the longer the stay, the higher the risk of weight loss. The average age of the patients in this study was higher, and the average length of hospital stay was longer than that of previous studies. Additionally, many patients in this study had cardiovascular disease (29.7%) and may have experienced edema on admission, which could have contributed to weight loss as the edema improved during hospitalization.

In this study, 15.9% of patients experienced a decline in walking ability during hospitalization. Valiani et al.^31)^ evaluated activity using the Braden Scale and reported that 20.6% of patients showed a decline in walking ability. Compared with the previous study, the patients in our study differed in several characteristics, including being originally independent in ADL and undergoing rehabilitation during hospitalization. The previous study reported a median hospital stay of 4 days, whereas in the present study it was 19 days. These differences may have contributed to the variation in the incidence of hospital-associated walking ability decline.

While most variables had low levels of missing data, variables involving body weight—such as GLIM-defined malnutrition, body weight before admission, weight on discharge, and weight loss during hospitalization—demonstrated comparatively higher missingness. These variables require accurate body weight measurement or reliable information on previous body weight; however, in routine clinical practice, obtaining such data is not always feasible. Furthermore, the need for multiple parameters to diagnose malnutrition according to the GLIM criteria may have increased the assessment burden and contributed to incomplete documentation in clinical practice. Missingness in GLIM-defined malnutrition was concentrated in specific facilities. This pattern may reflect differences in assessment systems or data entry processes across institutions. Although it may be challenging to mandate a full GLIM assessment, in Japan’s Diagnosis Procedure Combination system, body weight at both admission and discharge is already a required data item. Therefore, we are considering making body weight on discharge a mandatory variable in future database construction. To improve data completeness in future studies, standardization of required data elements and strengthening of centralized monitoring procedures will be necessary.

This study has several limitations. First, the number of participating facilities was limited, which may affect the representativeness of the patient population. Second, some evaluation indicators were based on simple screening tools and did not include detailed clinical diagnoses. Third, we did not systematically record the specific screening tools used prior to GLIM-based diagnosis. In addition, we did not establish a detailed protocol for body weight assessment, nor did we standardize the method for confirming previous body weight (3–6 months prior to admission). Therefore, concerns remain regarding the validity of the nutritional diagnosis and body weight–related data. Fourth, all patients included in this study were followed up until discharge. However, the total number of potentially eligible patients, the number screened, and the number excluded were not available. In addition, there was substantial variability in the number of registered patients across facilities, which may limit the generalizability of the findings. Some facilities had relatively small sample sizes, and direct comparison of incidence rates between facilities is limited by statistical instability. Therefore, it is difficult to draw definitive conclusions regarding interfacility differences in HAD incidence based on the present data, and the findings should be interpreted with caution. Fifth, it was difficult to obtain data on patients not referred for rehabilitation; therefore, the study population was limited to patients receiving rehabilitation, which may restrict generalizability. Although each participating facility was requested to enroll consecutive cases, adherence to this approach could not be monitored and may not have been fully ensured. In addition, patients without available preadmission ADL data were excluded, which may have reduced the sample size and introduced selection bias.

Nevertheless, this study, which gathered data prospectively from multiple centers, is significant for understanding the current status of HAD in clinical practice and for laying the groundwork for future analytical research.

This study elucidates the realities of HAD and its associated functional decline in an older adult inpatient population, providing a descriptive analysis of its characteristics, incidence, and background factors. In this study, 29.1% of patients experienced HAD, whereas 16.7% developed hospital-associated dysphagia. These findings are important for understanding the functional decline and risks experienced by older patients during hospitalization. The dataset obtained in this study may be utilized in future analyses to identify predictors of HAD and to develop early intervention programs for high-risk patients. The registry provides a structured foundation for future analytical studies.

## Data Availability

The datasets generated in this study are available from the corresponding authors upon request.

## Acknowledgments

This work was supported by JSPS KAKENHI (grant number 22K19669).

